# Trends in Lyme disease incidence in England, 2013 to 2024

**DOI:** 10.64898/2026.09.11.26362844

**Authors:** Janie Olver, Mark Gideon Burdon, Eva Emanuel, Ali Datoo, Verena FermorDunman, Miranda Ferguson, Jessica I’Anson, Roberto Vivancos, Christina Petridou, Amanda Semper

## Abstract

Official Lyme disease incidence figures for England are based on the number of cases with a laboratory-confirmed diagnosis from the national reference laboratory. However, these figures underestimate the true incidence of acute cases since patients can also be diagnosed clinically without the need for laboratory testing. The use of a multiplication factor has been proposed to estimate total Lyme disease incidence from the number of laboratory-confirmed cases. The current multiplication factor is derived from data spanning 1998 to 2016 and is thought to be outdated, in part due to the introduction of new national Lyme disease management guidance in 2018. The aim of this study was to analyse more recent laboratory and primary care data to identify trends in case numbers and to derive updated estimates of the total annual number of acute Lyme disease cases in England. Data spanning 2013 to 2024 from the national reference laboratory and a primary care data source were analysed using seasonal time series models. The presence of structural breaks was investigated using the PELT or BinSeg methods, and annual multiplication factors were estimated using a Poisson regression model. No substantial change was found in the number of laboratory-confirmed Lyme cases from 2013 to 2024 in England, whereas the number of cases diagnosed in primary care increased significantly over the same period, with an average of an additional 498 cases per year. This has increased our best estimate of the multiplication factor from 2.35 (95% confidence interval, 1.81 – 2.88) to 6.20 (6.17 – 6.24), which is the average for the years 2020 – 2024. Use of this updated multiplication factor should improve the accuracy of annual symptomatic Lyme disease case estimates for England.

## Introduction

Lyme disease is caused by pathogenic bacteria in the *Borrelia burgdorferi* sensu lato genospecies complex. In England, where it is the most common tick-borne disease, it is spread by the bite of infected *Ixodes ricinus* ticks [1].

The most common early presentation is erythema migrans (EM), an expanding skin rash at the site of the tick bite [1]. If untreated, disseminated infection can occur. In England the most common genospecies infecting ticks is *B. garinii* [2], which is particularly neurotropic. This is distinct from other European countries where the most common genospecies is B*. afzelii*, which primarily causes dermatological conditions. The third major pathogenic genospecies associated with Lyme disease is *B. burgdorferi* sensu stricto, which predominates in the United States and is particularly arthritogenic [1].

Clinical diagnosis can be made based on the recognition of the pathognomonic EM rash that is estimated to occur in 60-80% of infections in Europe [3]. In patients with non-specific symptoms, and no EM rash, serological testing is recommended. In England, this is performed using two-tier testing at the national reference laboratory, the Rare and Imported Pathogens Laboratory (RIPL), operated by the UK Health Security Agency (UKHSA) [4]. Samples can either be submitted directly or sent for confirmatory testing after testing positive (or indeterminate) locally.

Across Europe, various surveillance systems are used to estimate Lyme disease incidence, with estimates ranging from over 100 cases per 100, 000 people each year in Switzerland to less than 20 in Belgium [5]. However, caution is needed when interpreting these figures as surveillance systems differ in whether the data are collected regionally or nationally, and whether the case definitions use clinical or laboratory-confirmed cases, or both. In England, Lyme disease is not notifiable, but laboratory detection of the causative pathogen must be reported [6]. This means that the official incidence figures are based solely on the number of acute Lyme disease cases reported by the reference laboratory at UKHSA. These case numbers are published quarterly [7] and the resulting annual incidence figures are available online [8]. In 2024, for instance, the number of cases reported was 959, corresponding to 1.69 cases per 100, 000 people. However, this is known to underestimate the true incidence, as patients with an EM rash may be diagnosed solely in primary care without samples being sent for confirmatory testing.

Previous studies have compared the number of Lyme disease cases in primary care with the number of laboratory-confirmed cases, calculating a ‘multiplication factor’. This can be applied to the number of laboratory-confirmed cases to more accurately estimate the true incidence of symptomatic acute Lyme disease in England, enabling trends to be monitored over time.

The currently published multiplication factor is derived from a study that analysed primary care data from 1998 to 2016 [9]. The study used The Health Improvement Network (THIN) primary care dataset. This holds demographically-representative primary care data for 6.1% of the UK population, representing around 11.1 million patients [10]. The authors found that for every laboratory-confirmed case there were 2.35 (95% confidence interval, 1.81 – 2.88) cases identified in a primary care setting. Using this statistic, this means that in 2024, given the number of laboratory-confirmed cases reported online was 959 [7], the total number of symptomatic Lyme disease cases in England can be estimated to be 2, 254 (95% confidence interval, 1, 736 – 2, 762) corresponding to 3.8 cases per 100, 000 people (95% confidence interval, 3.0 – 4.7) [11].

A second study analysing Lyme disease cases diagnosed in primary care in the UK derived a higher incidence estimate ranging from 1.6 per 100, 000 people in 2001 to 12.1 in 2012 [12]. This study had a much broader case definition for Lyme disease, which may have led to an increase in the proportion of false positives.

It is suspected that the currently published multiplication factor of 2.35 may now be out of date in part due to the introduction in April 2018 of updated national guidance for the diagnosis and management of Lyme disease by the National Institute for Health and Care Excellence (NICE). This states that patients presenting with an EM rash should be diagnosed and treated without laboratory-confirmation [13]. The NICE guidance conforms with European recommendations [14].

This study aimed to provide an updated estimate of Lyme disease incidence in England through formal statistical analysis of trends in laboratory-confirmed cases and those diagnosed in primary care between 2013 and 2024. This has enabled the calculation of an updated multiplication factor, allowing more accurate estimation of the total number of annual symptomatic Lyme disease cases in England.

## Materials and Methods

### Data sources and case definitions

The study data included both UKHSA laboratory and National Health Service (NHS) primary care datasets.

### Laboratory-confirmed cases dataset

Official estimates of Lyme disease incidence in England are based on the number of patients who are determined by the reference laboratory to have acute Lyme disease. The definitions used for this have been described in detail in another report [15]. Further details on the assays used have also been described previously [16]. Case data are analysed for epidemiological reporting by UKHSA’s Emerging Infections and Zoonoses team (EIZ) to produce official yearly estimates of Lyme disease incidence in England [7]. This study used the annual epidemiological case data from 1^st^ January 2013 to 31^st^ December 2024 inclusive. Date of illness was taken as the sample collection date, if provided, otherwise the date of sample receipt at the reference laboratory was taken. The data had previously been filtered to only include patients whose residential postcode is in England.

### Sample submissions

Data from samples submitted to RIPL for Lyme testing between 1^st^ January 2013 and 31^st^ December 2024 inclusive were extracted from its laboratory information management system [17]. Prior to analysis, if samples were part of a CSF/serum pair, the CSF sample was removed, as were samples used for internal or external quality assurance purposes. Dates were derived using the same approach as for the laboratory-confirmed dataset. There were some samples where the date of receipt and the sample collection date were over two years apart. In these cases (95/192, 872 samples), as far as possible, the data were cleaned to rectify these data entry errors.

### Primary care dataset

The primary care dataset was extracted from the Clinical Practice Research Datalink (CPRD), specifically CPRD Aurum release September 2025, for events registered from 1^st^ January 2013 to 31^st^ December 2024 inclusive. The fields extracted were the patient, practice, and observation identifiers, as well as the date associated with the event, the date the event was entered into the system and the patient registration date. The codes for the medical term selected by the clinician (which are provided to the end user by CPRD as Read codes) and treatment were also extracted.

As recommended by CPRD, de-duplication at the practice and patient level was performed and practices that had merged with others during the study period were removed. Patient records were excluded if a consultation was entered before a patient’s registration date, as this would suggest they had moved practices. The data were filtered to only include English practices - no further geographical information on patients was available.

Data were extracted based on two criteria. The first was diagnosis with Lyme disease. This was defined as inclusion in the consultation of at least one Lyme disease-related Read code (Table S1). These Read codes were based on those used in previous studies [9, 12, 18]. The second was prescription of relevant antibiotics in the same consultation. Drug codes were chosen based on those recommended by the NICE guidance for treatment of Lyme disease (Table S2). Both the Lyme disease-related medical and antibiotic codes were reviewed by RIPL’s lead Lyme disease consultant.

As each patient can be assigned multiple Read codes per consultation, for instance erythema migrans (AA41-1) and Lyme disease (A8710), the data were restricted to the number of unique patient records with one or more Lyme disease-related codes. The date taken for time series analysis was the event date.

### Statistical analysis

All statistical analyses were carried out using R (version 4.3.2) and the underlying code is available on request. The code was independently reviewed by a principal infectious disease modeller within UKHSA.

Seasonal autoregressive integrated moving average (sARIMA) models were fitted to the sample submission, laboratory-confirmed, and CPRD primary care datasets using the auto.arima function available in the forecast library (version 8.23.0). This is because case (or sample submission) numbers are highly seasonal, being particularly high in the summer months, as this is when nymphal ticks, a key source of human infection, are most active in England [19]. Structural break analysis was undertaken to detect possible abrupt changes, using both the Pruned Exact Linear Time (PELT) and the binary segmentation (BinSeg) methods in the changepoint library (version 2.2.4) [20]. The statistical significance of these structural breaks was determined by comparing models with and without the structural break as an external regressor and comparing the difference in model fit (Akaike Information Criterion, AIC) to the Chi-squared distribution with one degree of freedom. All graphs were plotted using ggplot2 (version 3.5.1).

Annual acute Lyme disease incidence estimates per 100, 000 were calculated for the laboratory-confirmed data using the mid-year English population estimates for each year as the denominator population [11]. For the primary care data, the number of person years represented in CPRD was first calculated for each year following instructions available online [21] to enable calculation of the number of cases per 100, 000 people per year. Using these incidence estimates, the total number of estimated acute cases seen by GPs in England was derived for each year, using the same mid-year English population estimates as above [11].

Multiplication factors for each year in the study period, each with 95% confidence intervals, were estimated using a Poisson regression model. The average multiplication factors (overall, for 2013 – 2016 and for 2020 – 2024) were obtained by taking the arithmetic mean of the multiplication factor across included years. In this model, the number of cases was predicted by an interaction between individual year and the data source (CPRD primary care data or the laboratory-confirmed data) with offsets for the relevant population size. The data were checked for overdispersion. The 95% confidence intervals were estimated using the delta method [22]. The point estimates were equivalent and identical to manually dividing the primary care incidence by the laboratory-confirmed incidence.

## Results

### Trends in the number of laboratory-confirmed cases

From 1^st^ January 2013 to 31^st^ December 2024, there were a total of 10, 718 laboratory-confirmed cases. In 9% (n = 949) of cases, the sample collection date was not provided, so the sample receipt date was used instead as the date of illness. The number of monthly acute laboratory-confirmed Lyme cases is shown in Figure 1. The number of cases fluctuated throughout the year, with higher numbers of cases occurring during the summer months (June – September), as expected. The maximum number of cases diagnosed in a single month was 269 in August 2023, and the minimum number of cases was 8 (March 2014, April 2016 and February 2021).

**Fig One.**
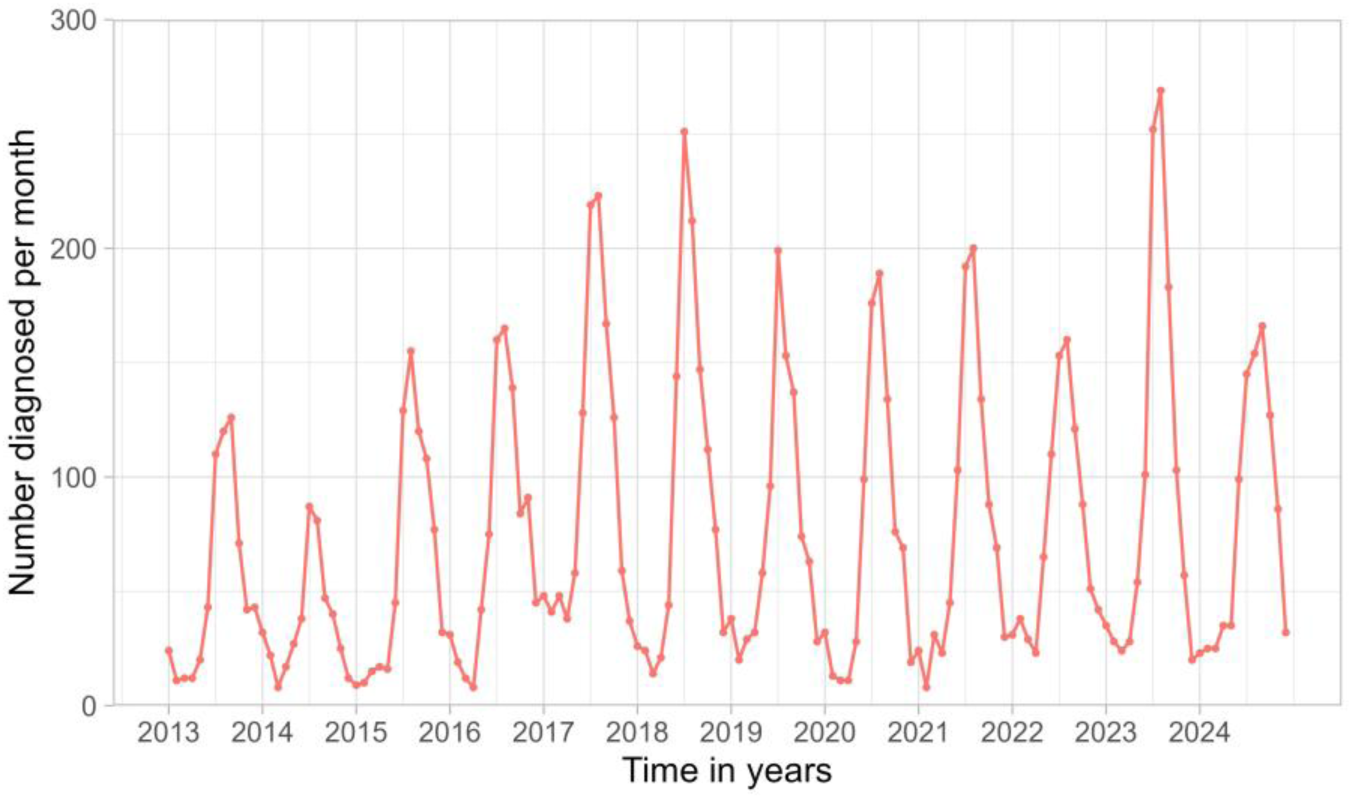
Monthly number of laboratory-confirmed acute Lyme disease cases in England, January 2013 to December 2024. Testing data from the reference laboratory (RIPL, UKHSA) cleaned for epidemiological reporting by UKHSA’s Emerging Infections and Zoonoses (EIZ) team.

Seasonal time series analysis revealed a drift coefficient over time of 0.0026 per month (standard error 0.0012), indicating a statistically significant increase over time. However, this corresponds to less than one additional case per year (0.0312 additional cases per year) and is therefore unlikely to be epidemiologically meaningful. No significant structural breaks were found between 2013 and 2024.

Although the number of laboratory-confirmed acute Lyme cases may have remained constant, the total number of samples submitted to the reference laboratory for Lyme disease testing may have changed. Therefore, trends in the number of samples submitted for Lyme disease testing were also analysed, using the sample collection date where available. This information was not given in 11% of cases (n = 21, 645), so date of receipt was used instead. The number of samples received by the reference laboratory per month is shown in Figure 2.

**Fig Two.**
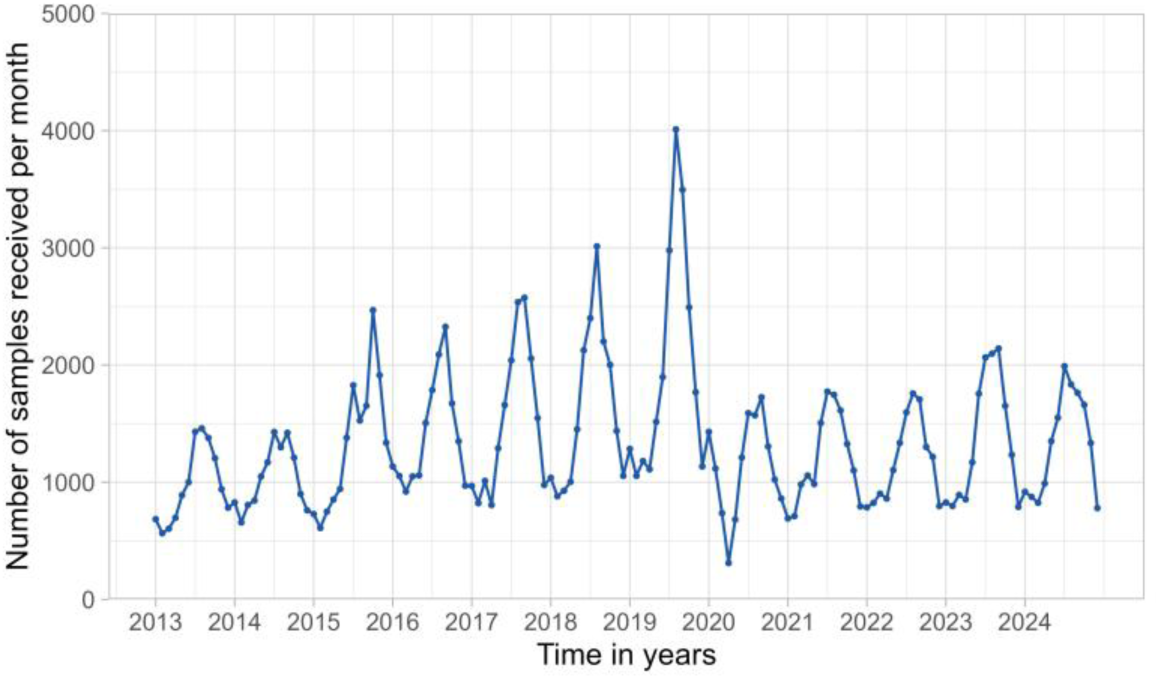
Monthly number of samples submitted to the reference laboratory for Lyme disease testing, January 2013 to December 2024.

As observed in the time series of the laboratory-confirmed case data, the number of samples fluctuated throughout the year, with submissions peaking during the summer months (June – September). The number of submitted samples reached its peak in August 2019, before dropping during 2020 and the COVID-19 pandemic. After 2020, the number of samples received remained at this lower level. Seasonal time series analysis did not identify a drift coefficient, and no significant structural breaks were found.

To investigate whether this drop in sample submission was due to particular healthcare providers ceasing to send samples to the reference laboratory, the share of total samples sent by each healthcare provider was analysed (Fig S1). In any year, no provider contributed more than 5% of the total samples received. Additionally, the median proportion of samples sent by any particular healthcare provider was approximately 0.2% of the total samples received per year.

Overall, there is no evidence of a significant increasing trend in the number of laboratory-confirmed Lyme disease cases in England, and the observed pattern is unlikely to be attributable to changes in the number of samples submitted.

### Trends in the number of cases diagnosed in primary care

From 1^st^ January 2013 to 31^st^ December 2024, there were a total of 15, 771 relevant primary care records in CPRD Aurum.

The usage of the different Lyme disease-related medical Read codes over time was analysed. A count of the number of times each code was used each year was calculated (Table S3). The most common code recorded over this study period was ‘Lyme disease’ (6, 792) followed by ‘Erythema migrans’ (3, 607). The rarest codes were ‘Acrodermatitis atrophicans chronica’, ‘Lyme carditis’ and ‘Lyme neuroborreliosis’, each with fewer than five occurrences.

The monthly number of acute Lyme cases presenting in primary care practices was then calculated and plotted (Fig 3A). The number of cases fluctuated throughout the year with peaks occurring during the summer months (May – October).

**Fig Three.**
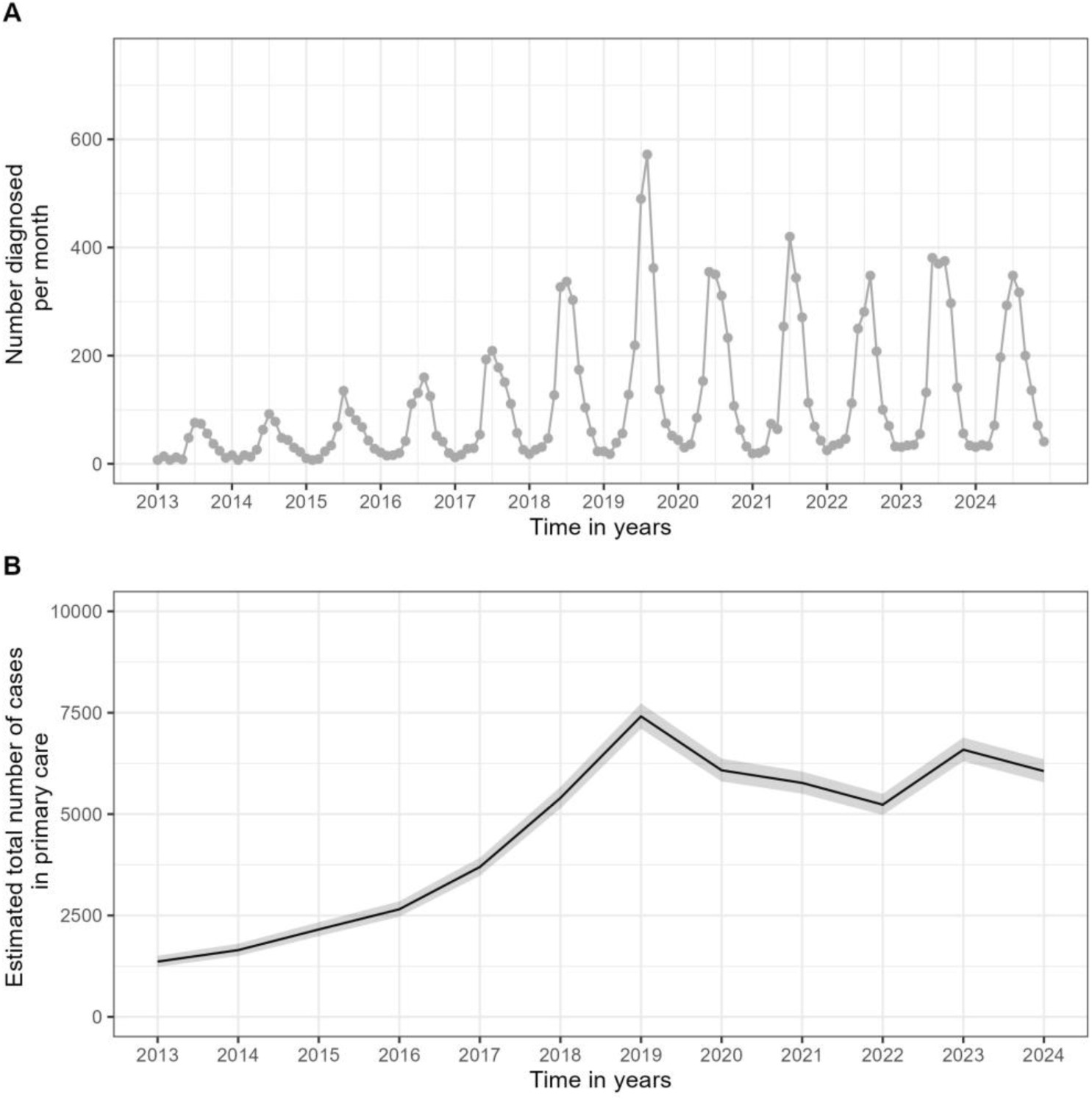
Lyme disease case numbers in primary care. (A) Monthly number of cases of acute Lyme disease in England identified in the CPRD Aurum dataset (release September 2025), January 2013 to December 2024. (B) Estimated total annual number of acute Lyme disease cases diagnosed in English primary care. The shading represents the 95% confidence intervals.

These numbers represent only the subset of patients whose primary care provider submits electronic patient health records to CPRD. This subset is geographically and demographically representative of the English population [23], and accounts for approximately 29% of the population [24]. To extrapolate case numbers to the entire English population, the CPRD denominator population and the English mid-year population estimate were used for each year to estimate the total number of Lyme disease cases diagnosed in primary care in England from 2013 to 2024 (Fig 3B and Table S4).

There was a significant increasing trend from 2013 to 2024, corresponding to an average increase of 498 acute Lyme disease cases diagnosed in primary care per year (standard error 94.6). The number of cases diagnosed in primary care peaked in 2019 at 7, 410 cases. From 2020 onwards, the trend appeared to stabilise at an average of a total of 5, 946 cases per year. No significant structural breaks were found between 2013 and 2024.

### Estimation of the annual incidence of acute Lyme disease cases in England

As expected from the significant increase in acute Lyme disease cases diagnosed within primary care, the multiplication factor significantly increased from 2013 to 2024 (Fig Four). This was used to estimate the annual number of Lyme disease cases in England from the number of laboratory-confirmed cases, as well as the corresponding incidence (Table One and Table S5).

**Fig Four.**
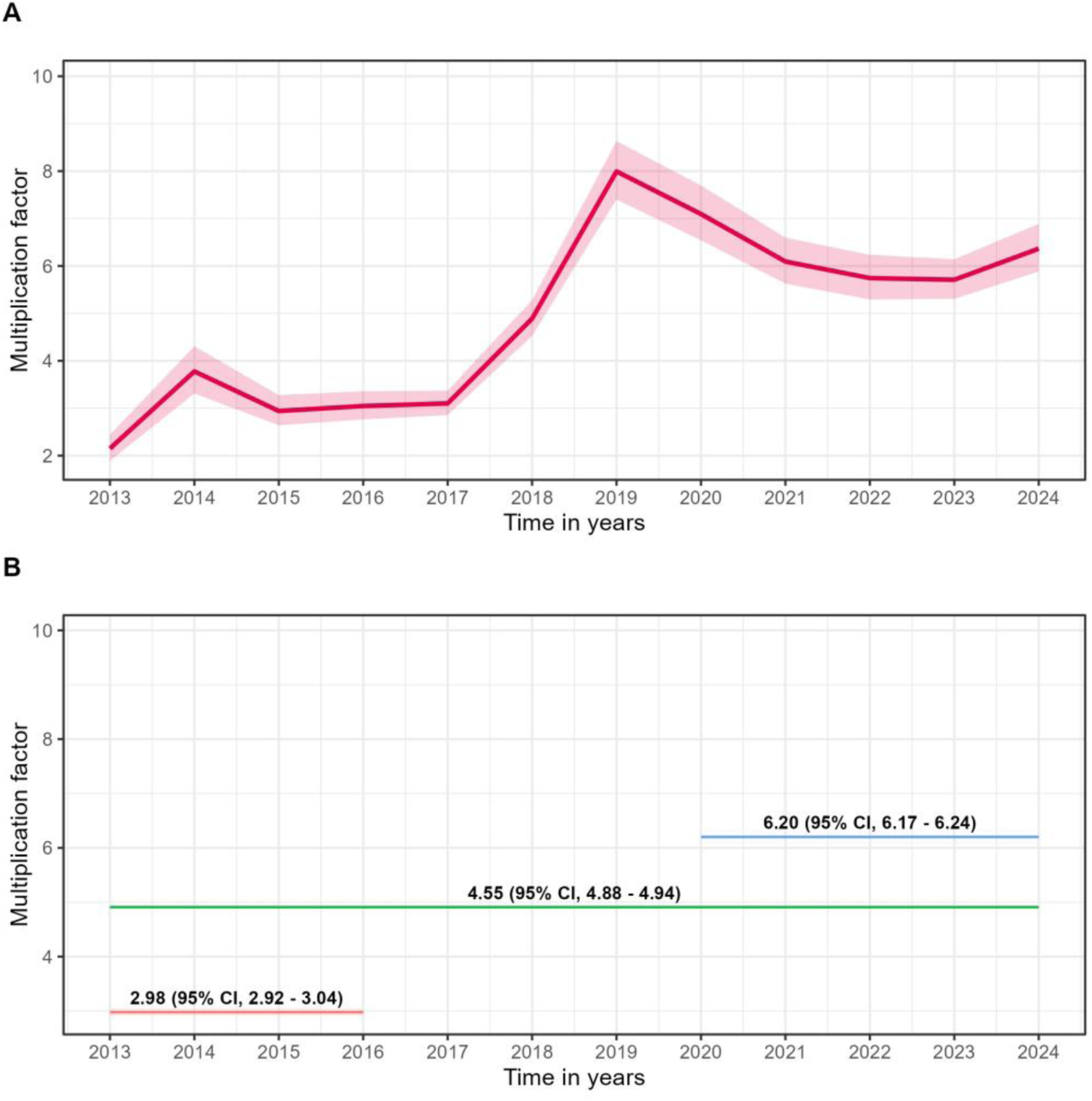
Multiplication factor estimates per year (A) and for specified year ranges (B). The multiplication factor for each year (A) with 95% confidence intervals (pale red), and the multiplication factor estimates with 95% confidence intervals for the specified year ranges for the specified year ranges (B).

**Table One.** Estimated annual case numbers and incidence of acute Lyme disease cases in England.

| *Year* | *Laboratory-confirmed cases* | *Multiplication Factor (95% CI)* | *Estimated total cases (95% CI)* | *Mid-year English population estimate* | *Estimated English incidence per 100,000 (95% CI)* |
| --- | --- | --- | --- | --- | --- |
| 2013 | 634 | 2.15 (1.89 – 2.44) | 1,363 (1,198 – 1,547) | 53,918,700 | 2.53 (2.22 – 2.87) |
| 2014 | 436 | 3.78 (3.31 – 4.31) | 1,648 (1,443 – 1,879) | 54,370,300 | 3.03 (2.65 – 3.46) |
| 2015 | 733 | 2.94 (2.64 – 3.28) | 2,155 (1,935 – 2,404) | 54,808,700 | 3.93 (3.53 – 4.39) |
| 2016 | 871 | 3.05 (2.76 – 3.36) | 2,657 (2,404 – 2,927) | 55,289,000 | 4.80 (4.35 – 5.29) |
| 2017 | 1,192 | 3.10 (2.86 – 3.37) | 3,695 (3,409 – 4,017) | 55,619,500 | 6.64 (6.13 – 7.22) |
| 2018 | 1,104 | 4.89 (4.53 – 5.28) | 5,399 (5,001 – 5,829) | 55,924,500 | 9.65 (8.94 – 10.42) |
| 2019 | 927 | 7.99 (7.40 – 8.63) | 7,407 (6,860 – 8,000) | 56,230,100 | 13.17 (12.20 – 14.23) |
| 2020 | 857 | 7.10 (6.54 – 7.70) | 6,085 (5,605 – 6,599) | 56,326,000 | 10.80 (9.95 – 11.72) |
| 2021 | 947 | 6.09 (5.63 – 6.60) | 5,767 (5,332 – 6,250) | 56,554,900 | 10.20 (9.43 – 11.05) |
| 2022 | 911 | 5.74 (5.29 – 6.23) | 5,229 (5,229 – 5,676) | 57,144,400 | 9.15 (8.43 – 9.93) |
| 2023 | 1,154 | 5.71 (5.31 – 6.14) | 6,589 (6,012 – 7,086) | 57,932,500 | 11.37 (10.58 – 12.23) |
| 2024 | 952 | 6.36 (5.88 – 6.89) | 6,055 (5,598 – 6,559) | 58,620,100 | 10.33 (9.55 – 11.19) |

*Values for the multiplication factor and incidence are given to two decimal places. The 95%* *confidence intervals are given in brackets. The mid-year English population estimate are those* *provided by the ONS [11].*

## Discussion

This study provides a statistical analysis of trends in acute Lyme disease cases in England between 2013 and 2024, comparing the number of laboratory-confirmed cases with the number diagnosed in primary care. Whereas the number of laboratory-confirmed cases has remained relatively constant from 2013 to 2024, the number of cases diagnosed within primary care has significantly increased. This has led to an increase in the multiplication factor, which can be applied to the annual number of laboratory-confirmed cases to calculate an estimate of the total incidence of acute Lyme disease in England per year.

No significant breakpoints were observed in the laboratory-confirmed, referral or primary care datasets around the time of the introduction of national NICE guidance on Lyme disease diagnosis and management in 2018. It is possible that its introduction did not produce a ‘step change’ but rather a gradual change which would not have been detected by the structural breakpoint estimation method. Indeed, the estimated number of cases diagnosed in primary care increased from 2018 to 2019, which could reflect increased clinical awareness of the NICE guidance. However, this seems unlikely, as there was already an increasing trend in the number of Lyme disease cases diagnosed in primary care from 2013 to 2018, which may have continued regardless into 2019. This increase prior to 2018 may have been due to increased public awareness, in part due to media reports of high-profile celebrity Lyme disease cases, increased awareness among health practitioners [25], or a true increase in Lyme disease incidence. This study cannot distinguish between these.

This study’s estimate of the overall number of acute Lyme disease cases in England per year is likely to be more representative than those derived in previous studies. This is because CPRD Aurum is not only geographically representative but is also representative of age and gender spread across England [23]. This means that this study’s extrapolation is likely to be more reliable than that of Cairns et al [12], who used an older version of CPRD which was not geographically representative [26] and for which post-hoc weighting of estimates was required. Similarly, the THIN dataset used by Tulloch et al does not guarantee geographical representativeness, particularly in the north of England, and they did not perform any post-hoc adjustments to their estimates to account for this [9]. It remains a limitation of this study, however, that it was not possible to stratify cases by demographic data, such as age, gender and geographical indicators like IMD scores and rural/urban scores. This is because these fields were not available for the laboratory-confirmed epidemiological data and are no longer available (and/or can no longer be derived from) data fields in CPRD Aurum.

This study aimed to reduce the number of false positives through its use of the dual criteria of both a recorded clinical diagnosis and prescription of appropriate antibiotics. However, it is not possible to ascertain how many of the patients included in the primary care analysis truly had Lyme disease. Some patients may have presented with a rash that was misidentified as erythema migrans and been treated unnecessarily or may have been given antibiotics simply as a precaution, contrary to national guidelines. Both may have been particularly true in 2020 and 2021 during the COVID-19 pandemic lockdowns in the UK, when patients were mostly seen via telephone consultation. Indeed, there is evidence for this in this dataset, with the use of ‘suspected’ codes increasing over two-fold for ‘Lyme disease’ and over six-fold for ‘erythema migrans’ when comparing pre-(2019) and pandemic (2020) Read code usage figures.

The combined usage of codes for ‘erythema migrans’, ‘erythema chronicum migrans’ and ‘suspected erythema migrans’ accounted for 40.30% of cases diagnosed in primary care in this study. This contrasts with Cairns et al [12] and Tulloch et al [9], who found that erythema migrans codes accounted for 27.97% and 21.85% of total code usage in each study respectively. It is likely that this study’s number is inflated by the ‘suspected erythema migrans’ code, which was introduced in 2014. This code either was not in use at the time of the previous study (Cairns et al, study period 2001 – 2012) or only used in the later years of the study period (Tulloch et al, study period 1998 – 2016), therefore only minimally contributing to the overall ‘erythema migrans’ figure. Excluding the ‘suspected erythema migrans’ code, this study observes a similar percentage of cases with an erythema migrans rash to Cairns et al and Tulloch et al (26.44%). It is therefore plausible that many of the patients coded with ‘suspected erythema migrans’ and treated are unlikely to be true cases. This means it seems likely that the primary care case numbers observed in this study are an overestimate of the true number of cases.

Another potential limitation is that no episode window was defined, meaning all unique primary care consultations meeting the dual criteria were included. This is different from Cairns et al [12] who only considered repeat consultations as separate episodes if they occurred more than one year apart. However, the choice of a year is rather arbitrary, particularly in an English context where there has been little study into the epidemiology of re-infection. Conversely, Tulloch et al [9] only included the first instance of a Lyme disease code for a patient and removed any subsequent instances. This was not done in this study as this would likely lead to an underestimation of cases, as it is physiologically feasible that some patients may have become re-infected during the time period of this study [27]. To avoid underestimation and the need to make potentially invalid assumptions, all unique consultations for each patient were included. A consultation was considered unique if it had a unique observation ID, observation date and date of entry onto the system. This may have led to overestimation of primary care case numbers and accordingly the estimate of total Lyme disease cases. Indeed, there were 65 patients with over three unique consultations recorded in CPRD Aurum over the period of the study. It seems unlikely that these are true re-infections, but this is also not implausible. In the absence of other data on these patients, such as laboratory results or photographs of the rash, this would be difficult to determine reliably.

Overestimation of the multiplication factor may also have occurred due to cases appearing in both the laboratory-confirmed and primary care datasets. This is because data on whether the laboratory-confirmed cases had an EM rash, and so were likely to have been diagnosed within primary care, was not available. Personally identifiable information was not available for either dataset, so it was not possible to link the datasets and remove such duplicates.

Underestimation may also have occurred due to missed cases. Cairns et al [12] were able to access the free text record within CPRD, which is no longer available in the version used here. Using text analysis of this field, they were able to identify additional Lyme disease cases. The cases that they identified through analysis of the free text records from 2001 to 2012 were overwhelmingly cases of suspected Lyme disease who were treated with antibiotics (1913/4083 of the total cases). In 2014, new ‘suspected’ Lyme disease Read codes were added, and so it is likely that such cases are no longer recorded in the free text field but captured using the more usual method of medical Read codes. This study would have identified these, as the ‘suspected’ Lyme disease codes were in the inclusion criteria. Indeed, in this study these codes were the third and fourth most used codes to describe Lyme disease with their usage increasing over time after their introduction in 2014. For instance, in 2015 ‘suspected Lyme disease’ accounted for 8.29% of the total number of Lyme-related codes used that year and this had increased to 25.89% in 2024. Similarly, ‘suspected erythema migrans’ accounted for 3.98% of codes in 2015 and this had increased to 22.45% in 2024. Therefore, despite the lack of the free text record in CPRD Aurum, it is unlikely that many cases of ‘suspected’ Lyme disease were missed in this study.

It is proposed that the current best estimate for total Lyme disease cases in England can be provided by applying the mean multiplication factor between 2020 and 2024, where it appears to have stabilised. This provides an average multiplication factor of 6.20 (95% confidence interval, 6.17 – 6.24). However, for this method to remain robust in the future, regular statistical analysis of Lyme disease case trends in primary care should be performed, because, as has been shown in this study, the number of cases diagnosed in primary care may change whilst the number of laboratory-confirmed cases remains relatively constant.

By using this updated average multiplication factor for 2020 to 2024 the estimate for the total number of Lyme disease cases in England in 2024 can be revised from 2, 254 (95% confidence interval, 1, 736 – 2, 762) to 5, 946 (95% confidence interval, 5, 917 – 5, 984), given that there were 959 laboratory-confirmed cases in 2024 [7]. This is an increase from an incidence of 3.8 cases per 100, 000 people (95% confidence interval, 3.0 – 4.7) to 10.1 cases per 100, 000 people (95% confidence interval, 10.1 – 10.2) [11].

Nonetheless, in comparison to other northern European countries, the English incidence remains very low. A recent study of primary care and other datasets in France suggests on average between 2017 to 2019 there were around 80 cases per 100, 000 people [28]. Similarly, a study in Germany that looked at healthcare data from 2015 to 2019 suggested that there were on average over 200 cases per 100, 000 people [29].

This study provides an updated estimate for the total number of acute Lyme disease cases in England, suggesting that this number has increased substantially than previously reported. Awareness of the increased estimate of total Lyme disease cases in England will enable more accurate data-driven improvements to be made to public health planning and action around Lyme disease risk in England.

## Acknowledgements

We would like to thank UKHSA members Emma Ackermann, Jonathon Mellor and Ayoub Saei for their help with both stylistic and statistical quality assurance.

## Statements

### Data availability

the primary care data were extracted from the Clinical Research Database Link (CPRD) and cannot be shared due to licensing restrictions. Applications for relevant anonymised laboratory-confirmed and referrals data should be submitted to the UKHSA Office for Data Release: Accessing UKHSA protected data - GOV.UK. The code is available on Git upon request.

### Ethical statement

this study is based in part on data from the Clinical Practice Research Datalink obtained under licence from the UK Medicines and Healthcare products Regulatory Agency. The data is provided by patients and collected by the NHS as part of their care and support. The interpretation and conclusions contained in this study are those of the authors alone. The approved study protocol number is 24_004190. No further ethical approval was required.

### Use of artificial intelligence tools

none declared.

### Contributions

Janie Olver (conceptualisation, data curation, formal analysis, validation, visualisation, writing – original draft, review and editing), Mark Gideon Burdon (formal analysis, validation, visualisation writing – review and editing), Eva Emanuel (data curation), Ali Datoo (data curation), Verena FermorDunman (data curation), Miranda Ferguson (data curation), Jessica l’Anson (data curation), Roberto Vivancos (conceptualisation, writing – review and editing, supervision), Christina Petridou (conceptualisation, writing – review and editing, supervision) and Amanda Semper (conceptualisation, writing – original draft, review and editing, supervision)

---

**Table S1.** – Lyme disease related Read codes.

| **Description** | **Read Code** |
| --- | --- |
| Lyme Disease | A8710 |
| Erythema Migrans | AA41-1 |
| Suspected Lyme Disease | 1JN1 |
| Erythema Chronicum Migrans | AA41 |
| Lyme Borreliosis | A8710-1 |
| Arthritis In Lyme Disease | N010A |
| Suspected Erythema Migrans | 1JN2 |
| Acrodermatitis Atrophicans Chronica | M21y0 |
| Lyme Arthritis | N010A1-1 |
| Lyme Neuroborreliosis | A8711 |
| Borrelia Lymphocytoma | A8713 |
| Lyme Carditis | A8712 |

**Table S2.**
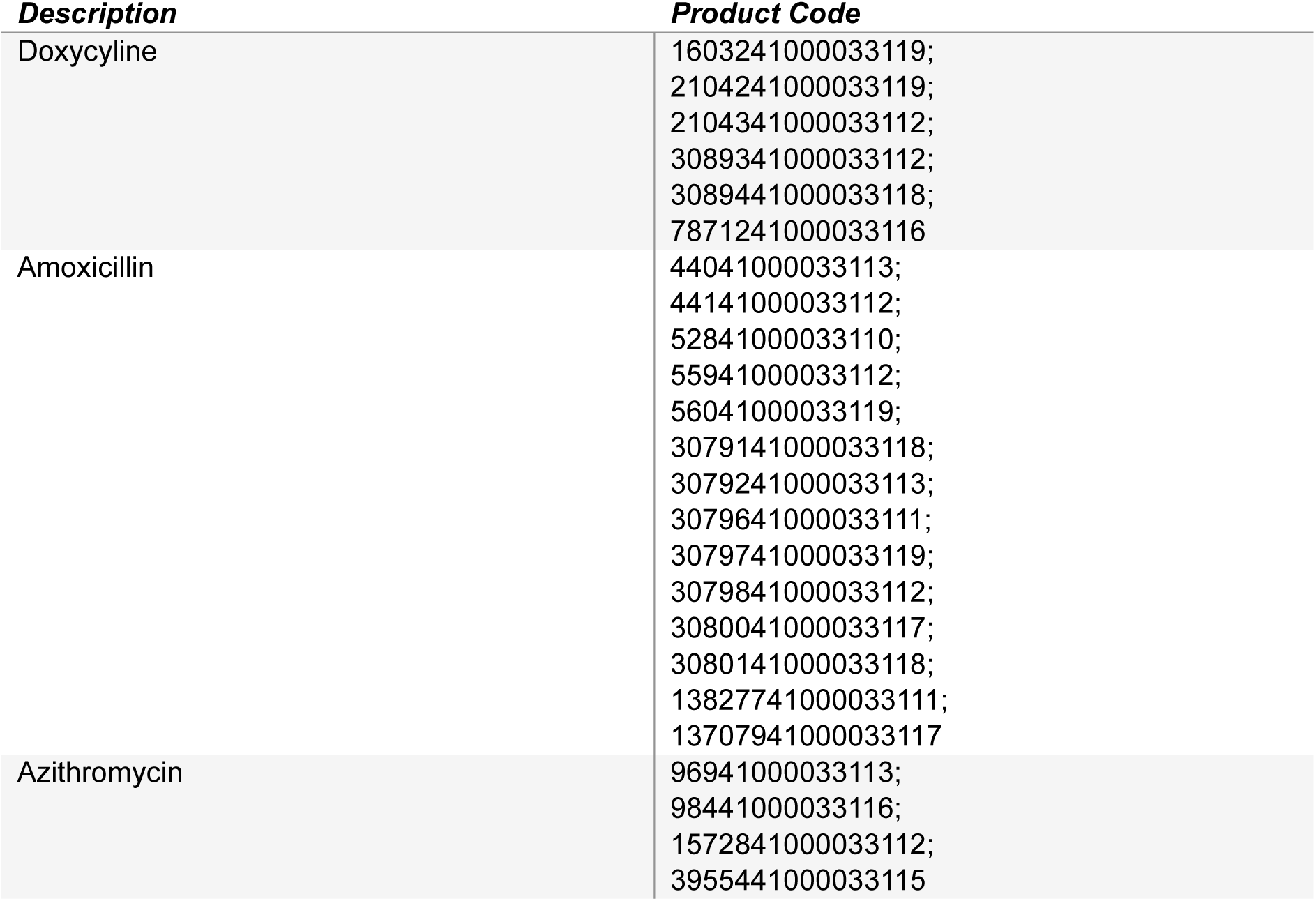
– Lyme disease related antibiotic codes.

**Fig S1.**
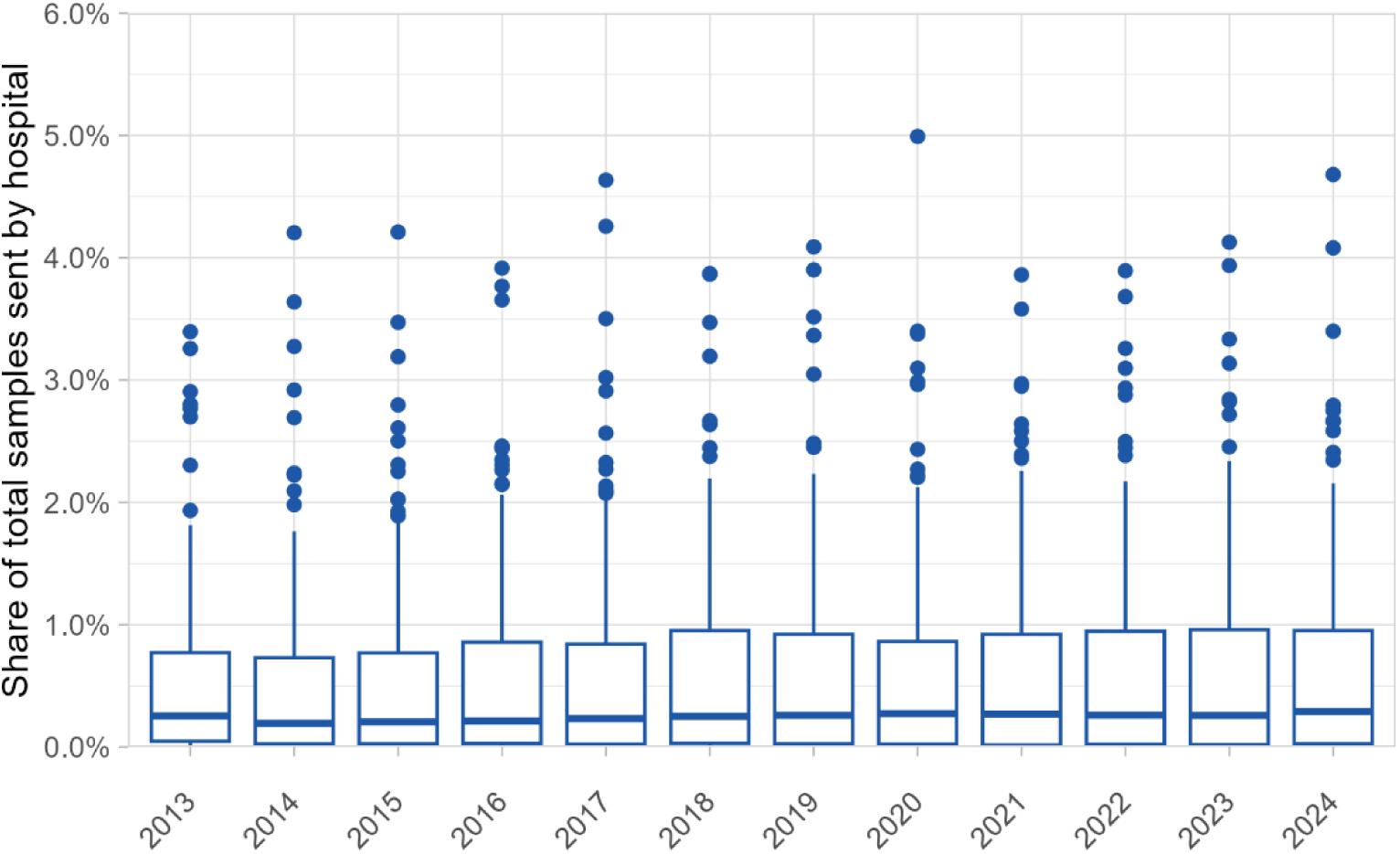
– Sample share per healthcare provider. The share of the total samples submitted by each healthcare provider per year. The lower and upper lines correspond to the first and third quartiles, and the middle line to the median. The ‘outlier’ points correspond to healthcare providers that send more than the value of 1.5x the interquartile range for each year.

**Table S3.** – usage of each Lyme disease related Read codes per year. Counts are provided (n) as well as the percentage (%). The percentage was calculated as the number of times each code was used in a particular year divided by the total number of recorded Read codes in CPRD Aurum for said year. The percentage is given to two decimal places. Codes that were used fewer than 50 times during the study period are not broken down by year. These are ‘acrodermatitis atrophicans chronica’ (n < 5), ‘arthritis in Lyme disease’ (n = 7), ‘Lyme arthritis’ (n = 12)), ‘Lyme borreliosis’ (n = 26), ‘Lyme carditis’ (n < 5) and ‘Lyme neuroborreliosis’ (n < 5). Similarly, the * indicates where values have been aggregated (combining both 2014 and 2015 yearly counts) or removed entirely to prevent re-identification.

| *Read Code Term* | **2013**<br>**(n)** | **2013**<br>**(%)** | **2014**<br>**(n)** | **2014**<br>**(%)** | **2015**<br>**(n)** | **2015**<br>**(%)** | **2016**<br>**(n)** | **2016**<br>**(%)** | **2017**<br>**(n)** | **2017**<br>**(%)** | **2018**<br>**(n)** | **2018**<br>**(%)** |
| --- | --- | --- | --- | --- | --- | --- | --- | --- | --- | --- | --- | --- |
| **ECM - Erythema chronicum migrans** | 87 | 23.26 | 94 | 20.66 | 53 | 8.79 | 45 | 5.97 | 46 | 4.32 | 79 | 5.01 |
| **Erythema migrans** | 0 | 0 | * | * | 113* | * | 168 | 22.28 | 243 | 22.82 | 507 | 32.17 |
| **Lyme disease** | 284 | 75.94 | 353 | 77.58 | 360 | 59.70 | 412 | 54.64 | 607 | 57.00 | 781 | 49.56 |
| **Suspected erythema migrans** | 0 | 0 | 0 | 0 | 24 | 3.98 | 42 | 5.57 | 39 | 3.66 | 52 | 3.30 |
| **Suspected Lyme disease** | 0 | 0 | * | * | 54* | * | 79 | 10.48 | 123 | 11.55 | 149 | 9.45 |
| **TOTAL** | 374 |  | * |  | * |  | 754 |  | 1065 |  | 1576 |  |

*Table S3 – usage of each Lyme disease related Read codes per year.
| *Read Code Term* | **2019**<br>**(N)** | **2019**<br>**(%)** | **2020**<br>**(n)** | **2020**<br>**(%)** | **2021**<br>**(n)** | **2021**<br>**(%)** | **2022**<br>**(n)** | **2022**<br>**(%)** | **2023**<br>**(n)** | **2023**<br>**(%)** | **2024**<br>**(n)** | **2024**<br>**(%)** | **TOTAL**<br>**2013 –**<br>**2024** |
| --- | --- | --- | --- | --- | --- | --- | --- | --- | --- | --- | --- | --- | --- |
| **ECM - Erythema chronicum migrans** | 57 | 2.63 | 22 | 1.22 | 28 | 1.63 | 16 | 1.04 | 18 | 0.93 | 17 | 0.96 | 562 |
| **Erythema migrans** | 798 | 36.76 | 371 | 20.62 | 405 | 23.60 | 307 | 19.90 | 367 | 18.91 | 328 | 18.50 | 3,607 |
| **Lyme disease** | 976 | 44.96 | 607 | 33.74 | 560 | 32.63 | 568 | 36.81 | 716 | 36.89 | 568 | 32.04 | 6,792 |
| **Suspected erythema migrans** | 77 | 3.55 | 420 | 23.35 | 426 | 24.83 | 330 | 21.39 | 379 | 19.53 | 398 | 22.45 | 2,187 |
| **Suspected Lyme disease** | 256 | 11.79 | 379 | 21.07 | 292 | 17.02 | 321 | 20.80 | 459 | 23.65 | 459 | 25.89 | 2,571 |
| **TOTAL** | 2171 |  | 1799 |  | 1716 |  | 1543 |  | 1941 |  | 1773 |  |  |

**Table S4.** – Calculation of total annual number of acute Lyme disease cases diagnosed in English primary care. Values for the total person years represented in CPRD are given as whole numbers. The mid-year English population estimates are those provided by the ONS [11].

| **Year** | **Cases in CPRD** | **Total person years represented in CPRD** | **Mid-year English population estimate** | **Estimated total number of cases in primary care** |
| --- | --- | --- | --- | --- |
| 2013 | 374 | 14,793,832 | 53,918,700 | 1,363 |
| 2014 | 455 | 15,025,221 | 54,370,300 | 1,646 |
| 2015 | 603 | 15,331,166 | 54,808,700 | 2,156 |
| 2016 | 754 | 15,710,765 | 55,289,000 | 2,653 |
| 2017 | 1,065 | 16,022,741 | 55,619,500 | 3,697 |
| 2018 | 1,576 | 16,327,300 | 55,924,500 | 5,398 |
| 2019 | 2,171 | 16,473,936 | 56,230,100 | 7,410 |
| 2020 | 1,799 | 16,664,771 | 56,326,000 | 6,081 |
| 2021 | 1,716 | 16,820,167 | 56,554,900 | 5,770 |
| 2022 | 1,543 | 16,849,422 | 57,144,400 | 5,233 |
| 2023 | 1,941 | 17,067,666 | 57,932,500 | 6,588 |
| 2024 | 1,773 | 17,153,188 | 58,620,100 | 6,059 |

**Table S5.** – Incidence Rate and Multiplication Factor Point Estimate for 2013 to 2024. The multiplication factor was calculated by dividing the incidence per 100, 000 in primary care (as estimated through CPRD Aurum) by the incidence of laboratory-confirmed cases. Values are given to two decimal places with 95% confidence intervals for each year’s multiplication factor in brackets.

| Year | *Estimated Incidence Rate per 100, 000 per Year* |  | *Multiplication Factor*<br>*(95% CI)* |
| --- | --- | --- | --- |
|  | *CPRD - Primary Care* | *Laboratory-Confirmed* |  |
| 2013 | 2.53 | 1.18 | 2.15 (1.89 – 2.44) |
| 2014 | 3.03 | 0.80 | 3.78 (3.31 – 4.31) |
| 2015 | 3.93 | 1.34 | 2.94 (2.64 – 3.28) |
| 2016 | 4.80 | 1.58 | 3.05 (2.76 – 3.36) |
| 2017 | 6.65 | 2.14 | 3.10 (2.86 – 3.37) |
| 2018 | 9.65 | 1.97 | 4.89 (4.53 – 5.28) |
| 2019 | 13.18 | 1.65 | 7.99 (7.40 – 8.63) |
| 2020 | 10.80 | 1.52 | 7.10 (6.54 – 7.70) |
| 2021 | 10.20 | 1.67 | 6.09 (5.63 – 6.60) |
| 2022 | 9.16 | 1.59 | 5.74 (5.29 – 6.23) |
| 2023 | 11.37 | 1.99 | 5.71 (5.31 – 6.14) |
| 2024 | 10.34 | 1.62 | 6.36 (5.88 – 6.89) |

